# CLOZAPINE-RELATED BRAIN *NRN1* EXPRESSION PATTERNS ARE ASSOCIATED WITH METHYLATION AND GENETIC VARIANTS IN SCHIZOPHRENIA

**DOI:** 10.1101/2024.10.29.24315982

**Authors:** Carmen Almodóvar-Payá, Marcos Moreno, Maria Guardiola-Ripoll, Mariona Latorre-Guardia, Benito Morentin, Beatriz Garcia-Ruíz, Edith Pomarol-Clotet, Luis F. Callado, Carme Gallego, Mar Fatjó-Vilas

## Abstract

The Neuritin-1 gene (*NRN1*), involved in neurodevelopment and synaptic plasticity, is associated with schizophrenia (SZ) and related clinical, cognitive, and neuroimaging phenotypes. Additionally, it is one of the most differentially methylated genes in the prefrontal cortex (PFC) in SZ and is responsive to neurotherapeutic agents. We aimed to investigate *NRN1*’s molecular mechanisms in SZ by analyzing its expression, methylation, and genotypic profiles in PFC and hippocampus (HIPP) post-mortem samples from 30 control (CTL) subjects and 20 individuals with SZ (10 treated with clozapine, SZ-Clz, and 10 without antipsychotic drugs at death, SZ-ApFree). We compared *the NRN1* mRNA expression between groups, measured by qPCR, and methylation levels across three CpG islands, assessed through EpiTYPER. Sparse Partial Least Square Discriminant Analysis identified key CpG units contributing to group differences. We then explored the relationship between *NRN1* methylation and expression, considering the influence of 11 polymorphisms genotyped by qPCR. We found that SZ-Clz had lower *NRN1* mRNA levels in the PFC than SZ-ApFree and CTL. SZ-Clz presented distinct methylation patterns across multiple CpG units in both brain regions compared to CTL. In the PFC, the methylation of the CpG units differentiating SZ-Clz from CTL correlated to *NRN1* expression, and the *NRN1*-rs12333117 and *NRN1*-rs2208870 polymorphisms influenced this effect. These findings reveal distinct correlations between *NRN1* epigenetic expression in SZ-Clz and CTL, shaped by genotypic variability. They emphasize region-specific alterations in SZ and underscore the importance of integrative approaches for a better understanding of the role of candidate genes in SZ etiology.

## 1. BACKGROUND

Despite schizophrenia (SZ) research progress, central pathophysiological mechanisms, molecular diagnostics, or precise biomarkers persist unclear. SZ is widely acknowledged to have a high ∼80% heritability (1,2), with a complex polygenic architecture involving numerous genetic variants (3). Large-scale genome-wide association studies (GWAS) have identified around 270 loci, many related to synaptic plasticity genes (4). However, these variants incompletely explain total heritability, with most not directly affecting protein structure but notably enriched for variants affecting DNA methylation, gene expression, and gene splicing in the human brain during different developmental stages (5). Together, these findings suggest that both genetic factors and epigenetic mechanisms regulating gene expression contribute to the developmental origins of SZ.

Among studies on epigenetic mechanisms, DNA methylation has been extensively researched, particularly in promoter regions for transcriptional repression, but it also regulates gene expression in distal elements like enhancers, exonic regions, and gene bodies, either inhibiting or activating expression based on genomic context (6). In the brain, methylation plays a twofold role: during development, the balance between methylation of germline-specific genes suppresses pluripotency, and demethylation of neuron-specific genes is key to facilitate neuronal specialization; in the mature brain, methylation changes can be induced by neuronal activity, contributing to complex cognitive processes such as learning and memory (7,8).

Methylation and gene expression studies in SZ, often performed utilizing prefrontal cortex (PFC) or hippocampus (HIPP) tissues, have shown distinct methylation patterns in genes critical for neuron development, synaptogenesis, and synaptic plasticity (9), as well as differential expression of genes related to immune response and inflammation, mitochondrial energy metabolism, myeloid leukocyte activation, cytoskeletal proteins, ion transport regulation, neurite outgrowth, and synaptic plasticity (10). Both types of studies have also suggested that these differences may be region and even cell-specific, with distinct genes implicated accordingly (11). Moreover, considering the impact of methylation on gene expression, numerous studies have correlated promoter hypomethylation of genes related to dopaminergic (*DRD2*, *DRD3*, *COMT*), GABAergic (*GAD1*), serotonergic (*HTR2A*), oligodendrocyte (*SOX10*), and synaptic plasticity (*RELN*) pathways in brain samples from patients with SZ with increased gene expression (12–15).

The above-mentioned studies collectively indicate that exploring methylation’s influence on expression in genes involved in brain development and synaptic plasticity might be key to understanding the biological underpinnings of SZ. The Neuritin1 gene (*NRN1*) exemplifies this, as it supports neuronal progenitors and differentiated neurons during early development (16), promotes the growth and stabilization of axonal and dendritic arbors, and facilitates synapse maturation in later development (17), while it also participates in adult brain synaptic plasticity, particularly in the PFC and the HIPP (18,19). Although *NRN1* has received less attention than traditional GWAS candidate genes, its chromosomal region is linked to an SZ subtype marked by cognitive deficits (20) and genetic association studies have related *NRN1* variants to the risk for SZ, cognitive deficits, early onset, and changes in brain structure and function (21–24). Additionally, two studies comparing patients with SZ to control subjects (CTL) have pinpointed *NRN1*, firstly, as one of the most differentially methylated genes in the PFC in SZ, and secondly, as one of the differentially expressed genes significantly contributing to patient differentiation according to transcriptomic clustering (25,26). Also, various cellular and animal models have shown that *NRN1* expression changes directly impact brain function and that antipsychotic treatments can induce such changes. In animal models, increased Nnr1 expression has been linked to better cognitive performance (27), improved recovery after ischemia-reperfusion injury (28), and protective effects in traumatic brain injury (29). Similarly, Nrn1 expression responds to neurotherapeutic agents like electroconvulsive therapy and fluoxetine through epigenetic pathways involving histone deacetylation (30,31). These findings suggest that modulating the *NRN1* expression could improve SZ symptoms and highlight its potential as a therapeutic target.

Among individuals with SZ undergoing antipsychotic therapy, special attention is given to those with a treatment-resistant profile, which affects about one-third of cases and significantly lowers their quality of life (32). Clozapine, the first atypical antipsychotic, has shown effectiveness in this group (33). It binds to various receptors (34), including dopamine, serotonin, histamine, muscarinic, and adrenergic receptors. Although the exact molecular pathways critical to its effectiveness are unclear, animal studies suggest that clozapine improves behavioral outcomes by modulating genes related to cholesterol metabolism, GABAergic function, cell cycle control, neurotrophins, and synaptic plasticity through reducing methylation or producing histone modifications (35). Meanwhile, human studies are constrained by limited access to post-mortem brain samples and the infrequent use of this treatment among patients. One study using publicly available SZ transcriptomic data found that three genes (*GCLM*, *ZNF652*, *GYPC*) and four pathways involved in protein trafficking, neuronal migration, brain development, and synaptic function were consistently differentially expressed in clozapine-treated patients compared to those on other medications (36).

Overall, current data highlights the role of genetic variability and gene expression regulatory mechanisms, such as methylation, in developmental abnormalities and the brain’s response to stimuli, contributing to SZ pathophysiology. Antipsychotics seem to modulate these epigenetic marks, suggesting new therapeutic targets. Therefore, this study aimed to investigate expression, methylation, and genotype variability of *NRN1*, a synaptic plasticity gene linked to SZ and sensitive to neurotherapeutic agents, in post-mortem PFC and HIPP samples from CTL and patients, considering antipsychotic treatment as a key factor.

## 2. METHODS

### 2.1. Post-mortem human brain samples description

Human post-mortem brain samples were collected at the Basque Institute of Legal Medicine (Bilbao, Spain), following research and ethics committee guidelines (Law 14/2007 and RD 1716/2011). It comprised 50 subjects, 20 with an ante-mortem diagnosis of SZ and 30 CTL. The diagnosis was performed by a qualified psychiatrist according to DSM-IV or ICD-10 criteria, as recorded in subjects’ medical records. Control subjects had no evidence of any psychiatric disorder in their medical records. Exclusion criteria for both groups included neurological conditions and substance abuse. Post-mortem brain samples from the dorsolateral PFC (Brodmann’s area 9) and the HIPP, excluding white matter, were dissected at the time of autopsy and immediately stored at −80°C until posterior analyses. Blood samples of those subjects were used for toxicological screening for antidepressants, antipsychotics, psychotropic drugs, and ethanol performed at the National Institute of Toxicology (Madrid, Spain). For CTL, selection ensured the absence of antidepressants or antipsychotic drugs at the time of death. Finally, both groups were selected to be matched by sex, age, and post-mortem interval (PMI), referring to the time elapsed between the death and the autopsy. Subjects with SZ were subsequently divided according to the presence of clozapine (SZ-Clz) or the absence of antipsychotic medication (SZ-ApFree) in the blood at the time of death. The summary of demographics and sample quality is shown in **Table 1** (see **Table S1** for individual details).

**Table 1.** Sample characteristics of control subjects (CTL) and individuals diagnosed with schizophrenia (SZ), categorized based on the presence of clozapine (SZ-Clz) or absence of antipsychotic medication (SZ-ApFree) in the blood at the time of death. Demographic information includes sex with female / male (F/M) count and frequency (%) for female category, age (in years), and cause of death. Assessment of post-mortem sample quality encompasses pH and post-mortem interval (PMI, in hours). All the quantitative variables are expressed as mean (standard deviation), while all the qualitative variables are presented as count (percentage). N=natural, O=others (including accidents and homicides), S=suicide.

| | CTL (n=30) | SZ-ApFree (n=10) | SZ-Clz (n=10) | $\chi^2$ / U | p-value |
| --- | --- | --- | --- | --- | --- |
| Sex (F/M) | 10 (33.3%) / 20 | 3 (30.0%) / 7 | 4 (40.0%) / 6 | 0.238 | 0.888 |
| Age | 44.03 (12.10) | 46.20 (10.14) | 48 (14.90) | 0.804 | 0.658 |
| Death cause (N/O/S) <sup>a</sup> | 11 (37%) / 19 (63%) / 0 | 5 (50%) / 3 (30%) / 2 | 4 (40%) / 1 (10%) / 5 | 2.397 | 0.302 |
| PMI | 14.13 (6.65) | 16.00 (4.81) | 19.20 (4.13) | 5.589 | 0.061 |
| pH | 6.37 (0.35) | 6.44 (0.28) | 6.38 (0.29) | 0.920 | 0.631 |

<sup>a</sup> This analysis compares SZ-Free with SZ-Clz, as CTL did not die by suicide according to the inclusion criteria.
<sup>b</sup> Data for pH was available for 27 CTL, 9 SZ-Clz and 6 SZ-ApFree.

### 2.2. Expression analyses

Total RNA was extracted using the NucleoSpin RNA Plus Kit (Cultek). RNA integrity and purity were analyzed by agarose gel electrophoresis and spectrophotometry using a NanoDrop. Reverse transcription reaction was performed with a maximum of 1µg of RNA using the Transcriptor First Strand cDNA Synthesis Kit (Roche) with random hexamer oligonucleotides. For quantitative PCR (qPCR) quantification, 1µL of the reverse transcription reaction was added to the qPCR reaction using JumpStart Taq ReadyMix for quantitative PCR (Sigma). qPCR reaction was preceded with an initial denaturation step at 94°C for 5 minutes followed by 45 cycles of: denaturation at 94°C for 15s, annealing at 51°C for 20s, and extension at 72°C for 30s. The fluorescent signal resulting from the degradation of the TaqMan probe (6xFAM/Black Hole Quencher1) was read at the end of the amplification phase. The primer sequences and probes are described in **Table S2**.

### 2.3. DNA extraction

Genomic DNA was extracted from post-mortem brain samples using the Quick-DNA Miniprep Plus Kit (Zymo Research) and used for methylation and genotyping analyses.

### 2.4. Methylation

Methylation analyses targeted the CpGs along the three CpG islands (CGI) spanning the *NRN1* gene (CGI1 chr6:5998917-5999730, CGI2 chr6:6002288-6004772 and CGI3 chr6:6006457-6006810, reference genome UCSC-GRC38/hg38). Samples were analyzed using the EpiTYPER method (Agena Bioscience) at the Centro Nacional de Genotipado (CEGEN, http://usc.es/cegen/). This method examines CpG sites within amplicons ranging from 200 to 600 base pairs, being especially well-suited for extensive endeavors aimed at investigating specific regions. In our case, six amplicons were designed using Agena’s EpiDesigner software to amplify these regions of interest (**Table S3**).

The methylation values can range between 0 and 1, being 0 the value corresponding to a 0% methylation state and 1 the value associated with a 100% methylated state. If CpG sites are too close together, their methylation levels cannot be determined independently, so the methylation status of a fragment containing multiple sites is measured, reflecting the average methylation of all CpG sites within that fragment. Accordingly, we will refer to CpG units, which can comprise one or more CpG sites. Each CpG unit was denoted numerically based on its 5’ to 3’ genomic position on the forward strand sequence.

The methylation quality control protocol implemented by CEGEN involved excluding units deviating by over 10% from the established commercial controls (HMc, 100% methylated, and LMc, 0% methylated), as well as excluding the samples with standard deviation variances between replicas exceeding 10%. Additionally, we eliminated units and subjects with more than 35% missing values and replaced outlier methylation values with the maximum for z-scores ≥ 3 and the minimum for z-scores ≤ -3. In all, we have analyzed 63 CpG units for the PFC and 57 CpG units for the HIPP along three CGIs within the *NRN1* region (see the details in **Figure 1** and **Table S4**).

**Figure 1.**
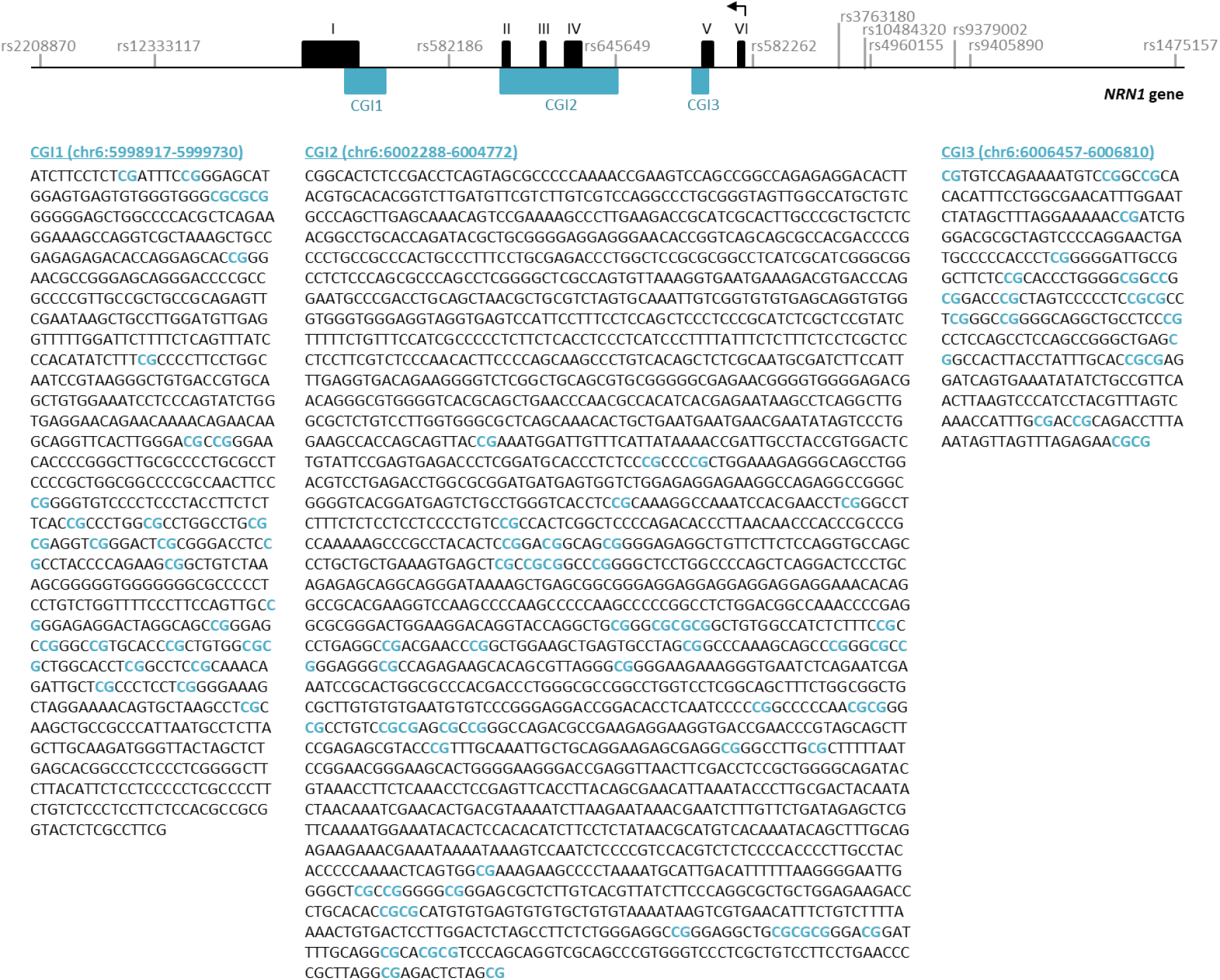
Schematic representation of the *NRN1* gene (NCBI36/hg18) with exonic (in black) and CpG island (CGI, in blue) regions, as well as single nucleotide polymorphisms (SNPs) included in the study according to the human genome browser (http://genome.ucsc.edu). Positions of the analyzed CpG sites in the DNA sequence are indicated below (in bold blue).

### 2.5. Genotyping

We genotyped eleven SNPs at the *NRN1* gene (6p25.1) at the National Genotyping Center (CEGEN, http://usc.es/cegen/) that were chosen to cover the genomic sequence and approximately 10 kb upstream and downstream. The optimal set of SNPs, which contained the maximum information about surrounding variants, was selected using the SYSNPs tool (http://www.sysnps.org/) with a minor allele frequency (MAF) > 5%, utilizing the pairwise tagging option (r² ≥ 0.8). Additionally, we included SNPs previously associated with SZ in the study by Chandler et al. (2010). All these SNPs have also been genotyped in other studies conducted by our group (21,21,23,37). The genotyping call rate was 100%, with minor allele frequencies closely matching those of the EUR population in the 1000 Genomes Project (**Table 2**). Genotype frequencies for both groups were in Hardy-Weinberg equilibrium (PLINK v.1.09, (38)).

**Table 2.** Information regarding *NRN1* SNPs included in this study. The table contains the dbSNP number, chromosome location (Chr Pos), alleles, and minor allele frequency (MAF) for each SNP based on the UCSC Genome Browser on Human (GRCh38/hg38). Additionally, MAF and genotypic count and frequency (%) are listed separately for control subjects (CTL) and subjects with schizophrenia (SZ), categorized based on the presence of clozapine (SZ-Clz) or absence of antipsychotic medication (SZ-ApFree) in the blood at the time of death. Due to low frequency in some genotypes, frequencies shown and subsequent analyses were conducted with all SNPs dichotomized (homozygous and heterozygous for the minor allele versus homozygous for the major allele).

| dbSNP | Chr Pos | Allele | MAF<sub>1000G</sub> | MAF<sub>sample</sub> | CTL freq | SZ-ApFree (n=10) | SZ-Clz (n=10) |
| --- | --- | --- | --- | --- | --- | --- | --- |
| rs2208870 | 6:5992257 | G/A | 0.34 | 0.33 | 16 (53.3%) / 14 | 4 (40.0%) / 6 | 6 (60.0%) / 4 |
| rs12333117 | 6:5994759 | T/C | 0.40 | 0.41 | 16 (53.3%) / 14 | 8 (80.0%) / 2 | 8 (80.0%) / 2 |
| rs582186 | 6:6001148 | A/G | 0.38 | 0.43 | 20 (66.7%) / 10 | 6 (60.0%) / 4 | 6 (60.0%) / 4 |
| rs645649 | 6:6004726 | C/G | 0.36 | 0.34 | 18 (60.0%) / 12 | 5 (50.0%) / 5 | 5 (50.0%) / 5 |
| rs582262 | 6:6007758 | C/G | 0.30 | 0.32 | 18 (60.0%) / 12 | 6 (60.0%) / 4 | 4 (40.0%) / 6 |
| rs3763180 | 6:6009615 | T/G | 0.46 | 0.48 | 22 (73.3%) / 8 | 7 (70.0%) / 3 | 7 (70.0%) / 3 |
| rs10484320 | 6:6010204 | T/C | 0.22 | 0.20 | 10 (33.3%) / 20 | 2 (20.0%) / 8 | 2 (20.0%) / 8 |
| rs4960155 | 6:6010306 | C/T | 0.49 | 0.52 | 22 (73.3%) / 8 | 8 (80.0%) / 2 | 8 (80.0%) / 2 |
| rs9379002 | 6:6012158 | G/T | 0.29 | 0.28 | 14 (46.7%) / 16 | 7 (70.0%) / 3 | 5 (50.0%) / 5 |
| rs9405890 | 6:6012488 | C/T | 0.31 | 0.26 | 12 (40.0%) / 18 | 3 (30.0%) / 7 | 4 (40.0%) / 6 |
| rs1475157 | 6:6016936 | G/A | 0.17 | 0.19 | 10 (33.3%) / 20 | 5 (50.0%) / 5 | 3 (30.0%) / 7 |

### 2.4. Statistical analysis

#### 2.4.1. Sample characteristics

For mean comparisons between cases and controls concerning sociodemographic and clinical characteristics as well as biological quality of the sample, Wilcoxon Rank Sum Test (for quantitative variables) and chi-square tests (for qualitative variables) were used.

#### 2.4.2. Expression analyses

Gene expression levels were determined with Delta Ct method (39), relativizing each gene expression to GAPDH levels. For the analysis, confounding variables (sex, age, death cause, PMI, and pH) were regressed from gene expression. Subsequently, a multifactorial ANOVA analysis was conducted followed by Tukey’s honest significance test, used for multiple comparisons of means between groups (CTL, SZ-ApFree and SZ-Clz).

#### 2.4.3. Methylation analyses

For methylation analyses, we assembled a subset consisting of 10 individuals each from the CTL, SZ-Clz, and SZ-ApFree groups, ensuring no significant differences with the initial sample in either demographic factors or post-mortem sample quality, as well as in *NRN1* expression across both regions (**Table S4** and **Figure S1**). Prior to analyzing methylation data, the confounding variables (sex, age, illness duration, and clozapine dosage) were regressed from each predictor methylation variable (CpG units).

To overcome the high intercorrelations among CpG units and the resulting multicollinearity (**Table S5**), we employed sparse Partial Least Squares-Discriminant Analysis (sPLS-DA), a method that simultaneously performs dimensionality reduction, feature selection, and classification to examine methylation differences between groups. Similar techniques, such as Principal Component Analysis (PCA), have been used in methylation studies to reduce high-dimensional genomic or locus-specific data into principal components that capture overall variance among CpG units in an unsupervised manner (40,41). In contrast, sPLS-DA is a supervised method that identifies latent components (LCs) by maximizing the covariance between the most relevant CpG units and group labels, enabling the discovery of methylation patterns associated with specific groups and enhancing the clinical significance of the findings.

All the analyses were based on the mixOmics protocol as implemented in the R package (42). To address the challenge posed by the small sample size in our study, we applied Leave-One-Out Cross-Validation (LOOCV) at multiple stages of the modelling process to calculate classification error. LOOCV is particularly well-suited to small datasets as it maximizes each data point’s use by treating each individual sample as a test set once, while training the model on the remaining data. First, LOOCV was used to determine the optimal number of LCs by evaluating the classification error across different component configurations. To further optimize the model, we applied the LASSO penalty in combination with LOOCV to select the most relevant CpG units. After determining the optimal number of LCs and CpG units, we used LOOCV to evaluate the performance of the final sPLS-DA model. We interpreted each CpG unit within an LC in two main ways: first, through its coefficient or loading value, which indicates its importance in distinguishing between groups, and second, by assessing the contribution of the CpG, which identifies the group that shows higher methylation values. Additionally, to validate the reproducibility of the CpG signature captured by the LCs, we performed a feature stability analysis, which assessed how frequently each CpG site was selected across multiple iterations of the model.

We selected the most informative LC using two criteria: i) first, following the sPLS-DA methodology, where the first component captures the maximum covariance between features and the outcome, which we further validated by comparing LC scores between groups using the Mann-Whitney U test; and ii) second, based on the stability of CpG units within each LC, as subsequent analyses focused on examining methylation patterns defined by these CpG units.

To further characterize the CpG signature captured by the LCs and find methylation patterns, following other studies with similar approaches (43,44), we computed two means: one grouping CpG units showing greater methylation in the SZ group (referred as “positive mean”); and another grouping CpG units with lower methylation in the SZ group (referred as “negative mean”). To validate the robustness of these patterns and ensure that the observed differences were not merely the result of a data-driven process, we also utilized the Mann-Whitney U test to statistically compare these means between the two groups.

#### 2.4.4. Expression-methylation correlates

We conducted four linear regression analyses using the expression residuals of the HIPP or the PFC as described in section *2.4.2* *Expression Analyses*. For each brain region, we tested two models: one with overall methylation variation (LC1) as the predictor and another with positive and negative mean methylation values as predictors.

#### 2.4.5. Genotypic analyses

We conducted four stepwise regression analyses using the expression residuals of the HIPP or the PFC as described in section *2.4.2* *Expression Analyses*. For each brain region, we tested two models: one with overall methylation variation (LC1) with 11 *NRN1* genetic variants (SNPs) as the predictors and another with positive and negative mean methylation values with 11 *NRN1* genetic variants (SNPs) as predictors.

## 3. RESULTS

### 3.1. Sample description

There were no differences in sociodemographic nor in the quality of the biological sample between groups, as they were matched for sex, age and PMI as shown in **Table 1**.

### 3.2. *NRN1* mRNA expression differences between groups

We found group differences in the PFC (F=10.160 and p=0.002) but not in the HIPP (F=2.576 and p=0.088). Tukey’s post-hoc analysis showed that SZ-Clz had significantly reduced *NRN1* mRNA expression levels in the PFC compared to CTL (Δ-means=-7.10 and p-adj=0.007) and SZ-ApFree (Δ-means=-8.85 and p-adj=0.005) (**Figure 2**).

**Figure 2.**
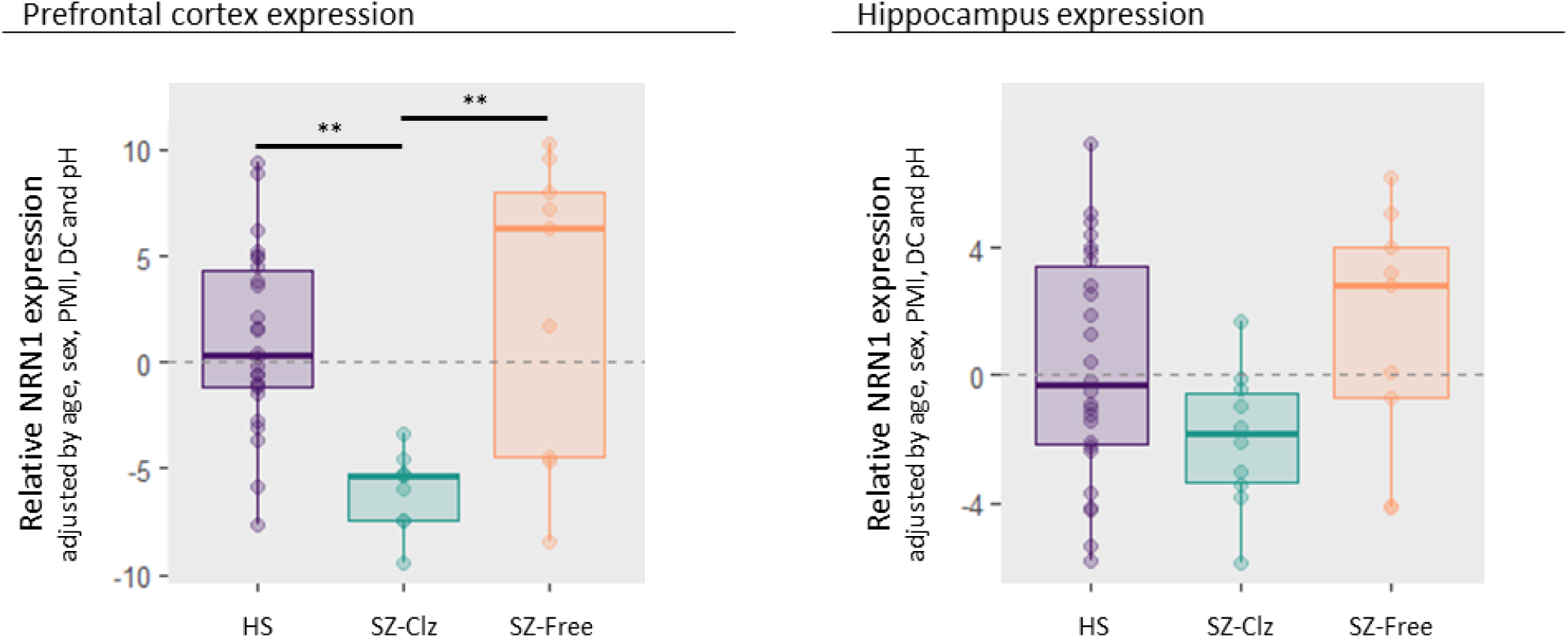
Boxplots representing NRN1 expression levels ± 2 standard errors (SE) depicted according to the group, control subjects (CTL), schizophrenia patients (SZ) categorized based on the presence of clozapine (SZ-Clz) or absence of antipsychotic medication (SZ-ApFree) in the blood at the time of death, for both brain regions: **A)** Prefrontal cortex (n=27, 9, and 9 respectively), and **B)** hippocampus (n=26, 9, and 10 respectively). The NRN1 expression was relativized to GAPDH, and the values expressed are the residuals (adjusted by sex, age, post-mortem interval (PMI), death cause (DC) and pH). Adjusted p-values from Tukey’s test are represented as * p<0.05, ** p<0.01, *** p<0.001.

To validate the findings concerning *NRN1* expression we also analyzed the expression of two other synapse-related genes well described to be sensitive to clozapine treatment, *UMHK1* and *BDNF* (45). Like *NRN1*, significant group differences emerged within the PFC for both *UMHK1* (F=5.73, p=0.006) and *BDNF* (F=8.993, p<0.001), while not in the HIPP. Tukey’s post-hoc analysis revealed that clozapine treatment was markedly associated with a decrease of *UMHK1* mRNA expression levels in the PFC compared to both CTL (Δ-means=-2.589, p-adj=0.032) and SZ-ApFree (Δ-means=-3.968, p-adj=0.005), while *BDNF* expression levels were notably higher compared to CTL (Δ-means=1.307, p-adj < 0.001) (**Figure S2**).

### 3.3. *NRN1* methylation differences between groups

Initially, our analysis encompassed all three groups, but we observed a limited ability to identify a specific linear combination of CpG units from either the PFC or the HIPP that could effectively differentiate between the three groups (balanced error for the model incorporating CpG units from the PFC=0.74 and from the HIPP=0.66). Furthermore, upon projecting the samples into the two LC spaces determined by the respective models, we observed a significant overlap between CTL and SZ-ApFree, but a clear distinction of SZ-Clz in both regions (**Figure S3 and Figure S4**).

Subsequently, we generated two separate models for each region, focusing on the CTL and SZ-Clz groups. In the PFC, three LCs explained 12%, 17%, and 10% of the methylation variance, respectively, effectively distinguishing the groups (balanced error=0.28) (**Figure 3A**). Similarly, in the HIPP, two LCs explained 34% and 22% of the variance, achieving the same level of separation (balanced error=0.28) (**Figure 3C**). Due to the sPLS-DA methodology, LC1 was the most influential in differentiating the groups in both regions. This was further confirmed by significant differences observed in PFC-LC1 (W=76, p<0.001) and HIPP-LC1 (W=71, p=0.005), while the other LCs did not reach statistical significance, neither in the PFC (LC2: W=62, p=0.06; LC3: W=51, p=0.38; **Figure 3B**) nor in the HIPP (LC2: W=61, p=0.08; **Figure 3D**). Additionally, PFC-LC1 and HIPP-LC1 exhibited greater feature stability, with at least 88% of CpG units showing high stability (>85%), compared to less than 50% in the other LCs (**Table S6**). Accordingly, subsequent analyses of methylation patterns among the CpG units focused on LC1 in both regions.

**Figure 3.**
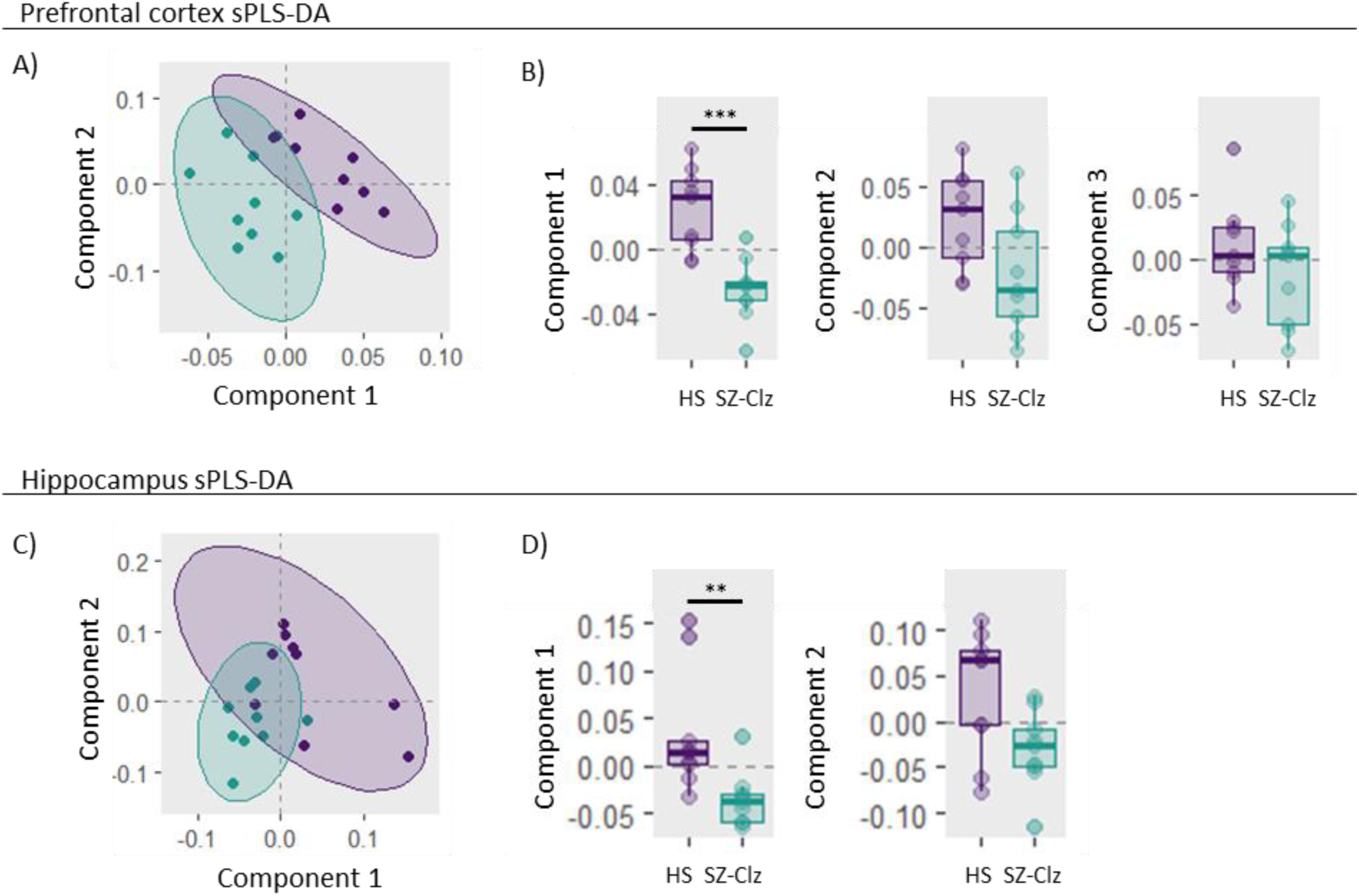
**A and C)** Scatter plot illustrating the distribution of samples projected into the space defined by the first two components derived from sparse Partial Least Square Discriminant Analysis (sPLS-DA) using *NRN1* methylation values from the hippocampus (HIPP). Each sample class is enclosed within 95% confidence ellipses. **B and D)** Boxplots representing methylation scores for the first and second latent components (LC) ± 2 standard errors (SE) depicted according to the group. Control subjects (CTL) are depicted in purple while schizophrenia patients treated with clozapine (SZ-Clz) are shown in blue. * p<0.05, ** p<0.01, *** p<0.001.

In both brain regions, LC1 included 10 CpG units with varying importance in distinguishing between classes. CpG units with positive weights, represented by red bars, exhibited higher methylation levels in SZ-Clz subjects compared to CTL, while those with negative weights, shown in green, exhibited lower methylation levels in SZ-Clz subjects (**Figure 4A and C**). In both regions, the mean of the CpG units in LC1 with reduced methylation in SZ-Clz subjects compared to CTL significantly differed between groups ("PFC negative average": CpG3.8, CpG2.5, CpG1.20, CpG2.6, CpG1.3, and CpG2.28, W=69, p=0.01; "HIPP negative average": CpG2.1, CpG2.2, CpG2.3, CpG2.5, and CpG3.3, W=70, p=0.008), suggesting that the observed differences are not merely data-driven artifacts. In contrast, the mean of the CpG units in LC1 with higher methylation in SZ-Clz subjects compared to CTL did not show significant differences in either region ("PFC positive average": CpG3.12, CpG1.2, CpG1.1, and CpG1.16, W=18, p=0.05, **Figure 4B**; "HIPP positive average": CpG1.19, CpG3.1, CpG3.4, CpG2.21, and CpG1.20, W=31, p=0.44, **Figure 4D**).

**Figure 4.**
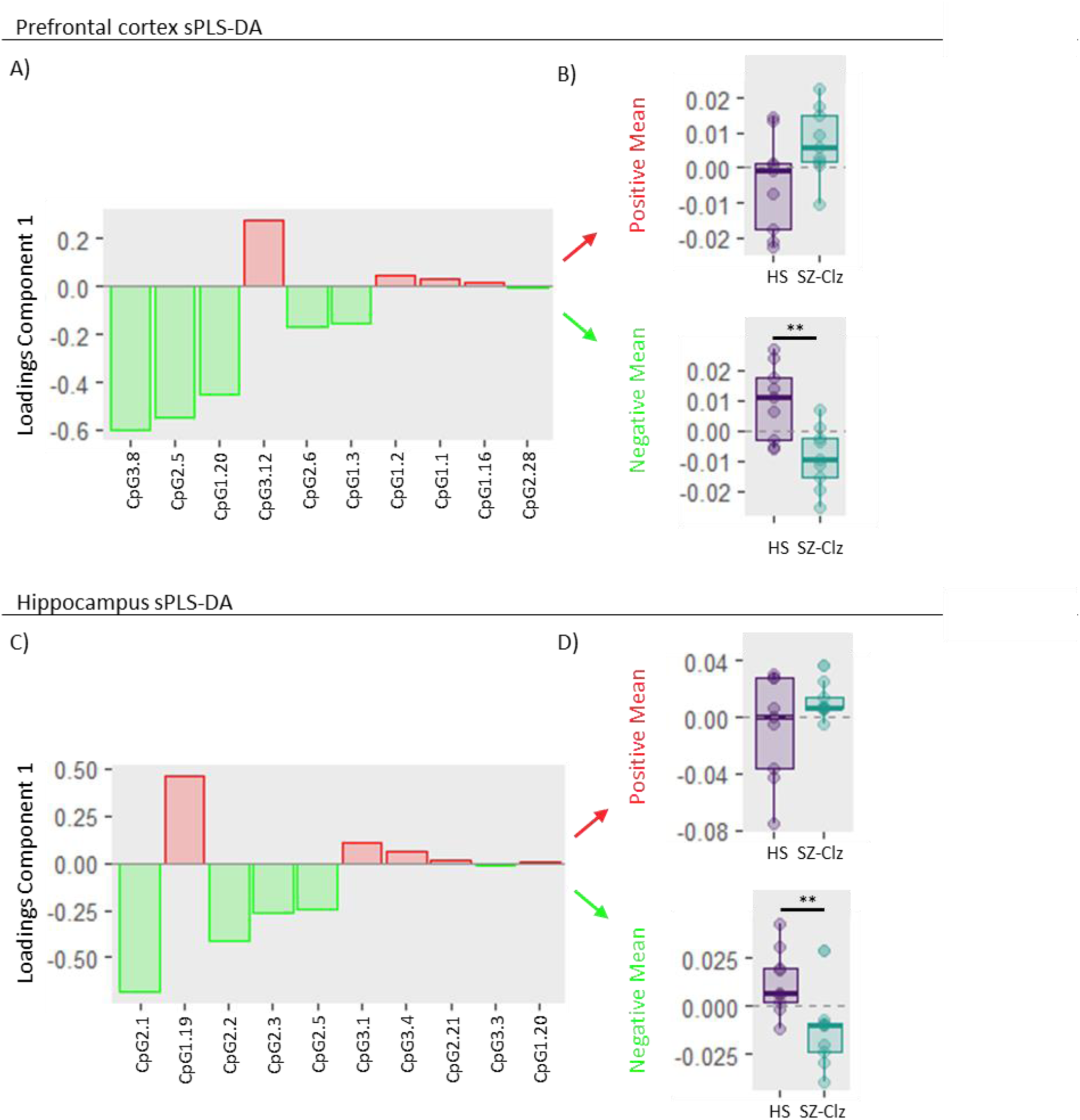
**A and C)** Bar plots illustrating the loading weights for each CpG unit in component 1. Red bars indicate higher methylation in patients with schizophrenia (SZ) treated with clozapine (SZ-Clz), while green bars represent lower methylation in SZ-Clz compared to control subjects (CTL). **B and D)** Boxplots representing the mean methylation values, computed based on whether the CpGs exhibited higher (positive mean) or lower (negative mean) methylation levels in SZ-Clz compared to CTL, ± 2 standard errors (SE) depicted according to the group. SZ-Clz are shown in blue and CTL in purple. * p<0.05, ** p<0.01, *** p<0.001.

### 3.3. *NRN1* expression methylation correlates

We observed a significant correlation between LC1 scores derived from the PFC sPLS-DA model and *NRN1* gene expression in the corresponding brain region across the sample including both SZ-Clz and CTL groups (standardized β=0.753, p<0.001, adj-R²=0.536). When we further analyzed the CpG units based on their methylation status in the SZ-Clz group, the correlation remained significant for the "negative mean" values (standardized β=0.715, p=0.002, adj-R²=0.477), while the "positive mean" values were not associated with NRN1 expression. These results indicate that lower LC1 scores or "negative mean" values are linked to a reduced NRN1 expression (**Figure 5** and **Figure S5**). Conversely, we did not find any correlation between NRN1 expression and methylation in the HIPP.

**Figure 5.**
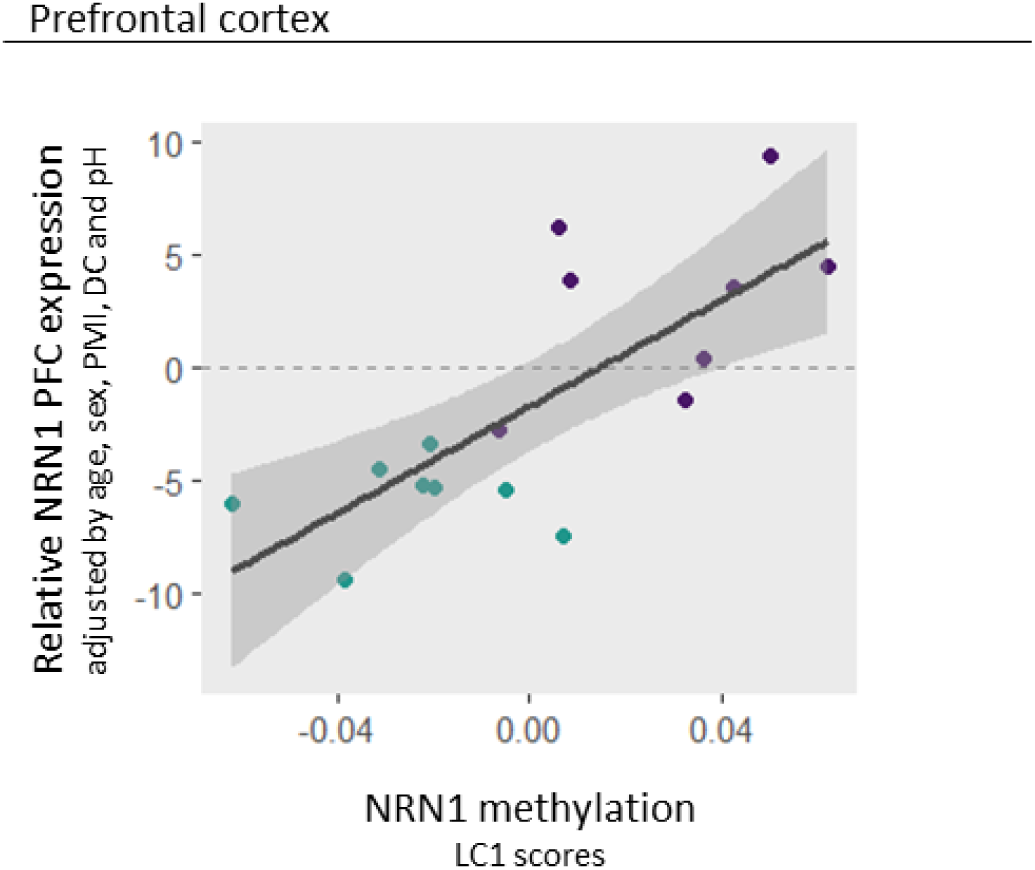
Scatter plot illustrating the correlation between latent component 1 (LC1) scores derived from the sparse Partial Least Square Discriminant Analysis (sPLS-DA) of the prefrontal cortex (PFC) and *NRN1* gene expression levels in the same region. *NRN1* gene expression levels are shown with ±2 standard errors (SE). The expression of *NRN1* was normalized to GAPDH, and the values are presented as residuals, adjusted for sex, age, post-mortem interval (PMI), cause of death (DC), and pH. The sample includes both schizophrenia patients treated with clozapine (SZ-Clz) and control subjects (CTL), with dots colored blue for SZ-Clz patients and purple for CTL.

### 3.4 *NRN1* genetic variability effects

We found that incorporating two genetic variants, rs2208870 and rs12333117, along with LC1 scores or the "negative mean" values from the PFC sPLS-DA model, enhanced the correlation with *NRN1* gene expression in the corresponding brain region in the sample including SZ-Clz and CTL groups. Adding LC1 scores alongside the two genetic variants significantly enhanced the model’s performance compared to using only the methylation variable (model’s F=11.3, Adj-R^2^=0.673, p=0.001; Δ-R^2^=0.137, p of the change=0.048). LC1 scores exhibited a robust association with NRN1 gene expression (standardized β=0.770, p<0.001), while the genetic variants also made significant contributions (standardized β=-0.404, p=0.029 for rs2208870; standardized β=-0.364, p=0.045 for rs12333117). Similar improvements were observed when incorporating the "negative mean" values alongside the two genetic variants, significantly enhancing the model’s performance compared to using only the methylation variable (model’s F=12.52, Adj-R^2^=0.697, p=0.001; Δ-R^2^=0.221, p of the change=0.015). The "negative mean" exhibited a strong association with NRN1 gene expression (standardized β=0.790, p<0.001), while the genetic variants also made notable contributions (standardized β=-0.490, p=0.010 for rs2208870; standardized β=-0.442, p=0.016 for rs12333117). Conversely, we found that incorporating genotypic variability into the methylation metric did not enhance the model’s ability to explain *NRN1* expression in the HIPP.

## 4. Discussion

Given the crucial roles of *NRN1* in brain development and synaptic plasticity, and the significance of these mechanisms in the etiology of SZ, we have investigated the molecular variability of this gene in post-mortem brain samples from patients with SZ and CTL. Our study reveals that *NRN1* is significantly under-expressed in the PFC of patients treated with clozapine and that methylation levels correlate with *NRN1* expression in the same region. In addition, our study shows that incorporating genetic variability data of *NRN1* further shapes this correlation, highlighting the necessity of multi-level approaches to understand the role of candidate genes in SZ etiology.

First, our results showed that patients treated with clozapine exhibited reduced expression of *NRN1* in the PFC as compared to both CTL and Sz-ApFree patients. On the one hand, these differences should be considered in terms of the their functional impact, as *NRN1* expression is crucial as it enhances synaptic transmission by increasing the surface expression of CaV1.2, CaV1.3, and CaV3.3 channels, mediated through the insulin receptor and ERK signaling pathways (46–48). Then, our data provides new evidence suggesting that the reduced NRN1 PFC expression observed in patients with SZ may be specifically associated with, or at least more pronounced under, clozapine treatment, and that such differences could be related to synaptic transmission variability. Despite the scarcity of comparable studies, to further interpretating our findings it is interesting to mention that epigenetic mechanisms have been implicated in the effects of atypical antipsychotic treatment, specifically demonstrating that chronic treatment induces the expression of proteins related to histones deacetylation, such as HDAC2 (49). Particularly, clozapine treatment has been related to widespread promoter demethylation (50). Also, it is remarkable a previous study exploring dorsolateral PFC transcriptome of schizophrenia patients, that identified two molecularly distinct subgroups of patients, one similar to CTL and the other markedly different, with NRN1 being among the down-regulated genes between the latter subgroup and CTL (25). Then, we tried to replicate our results with a larger sample of clozapine-treated patients, but public data was limited and had fewer treated patients than our study (GSE224683, GSE80655, GSE12649). On the other hand, the fact that our study did not reveal the same effect in HIPP, is aligned with previous research showing different anatomical methylation signatures in patients with SZ and CTL when comparing the PFC and the HIPP, possibly due to varying cellular compositions and their corresponding functional specializations (51). This interpretation within the framework of region-specific transcriptome alterations in SZ mirrors the loss-of-function mutations in the *GRIN2A* gene, encoding an NMDA receptor subunit, which significantly increase SZ risk by differentially affecting brain regions and circuits (52,53).

Second, we explored the methylation profile of *NRN1* in the two brain regions, but recognizing the complexity of interpreting methylation changes in SZ, in a complementary approach to the global perspective given by the PLS, we advanced to more detailed interpretations to integrate both broad methylation patterns and targeted site-specific analyses. As regards the differences between SZ-Clz and CTL, defined by distinct CpG units in each region, we observed that the component LC1, which best distinguished the two groups, included CpG units with both increased and decreased methylation in patients, indicating that bidirectional changes are a defining feature of the patient methylation profile across both brain regions. However, upon grouping CpG units by the direction of methylation change, we found that the average of those CpG units under-methylated in SZ-Clz was significantly different between groups in both regions, suggesting that lower methylation in SZ-Clz may play a more prominent role. Notably, a methylome-wide study identified a large region (chr6:59992669-6006917) encompassing 29 adjacent CpG sites along the gene body of the *NRN1* as hypomethylated in PFC of SZ patients compared to CTL (26). This region included all CpG sites detected as hypomethylated in our study except for CpG 1.3, hence our findings add to those from Pidsley et al., 2014 while also suggesting that the observed hypomethylation may be characteristic of clozapine-treated patients.

Third, when exploring the relationship between *NRN1* methylation and expression patterns, we observed that two methylation metrics derived from the PFC model, LC1 and the average of those CpG units under-methylated correlated with *NRN1* expression in this region. Although the novelty of our study limits direct comparison with other *NRN1*-specific results, our findings are consistent with previous research linking DNA methylation to differential gene expression in post-mortem brain samples from individuals with SZ (54). Additionally, it is important to note that some studies have linked clozapine treatment, but not haloperidol or olanzapine, to changes in *BDNF* expression, an upstream regulator of *NRN1*, through the reduction of its promoter methylation (55).

Next, to gain further insights into the potential methylation-expression correlates, we identified regulatory elements overlapping with CpG units within PFC LC1 by using the UCSC Genome Browser (http://genome.ucsc.edu) to access data from the Encyclopedia of DNA Elements (ENCODE) project (56) and the Open Regulatory Annotation (ORegAnno) database (57). The presence of enhancer regions, DNase I hypersensitive sites within these CpG units, and histone marks like H3K27me3, H3K4me1, and H3K36me3 highlights their potential in gene expression control and chromatin accessibility. Upon examining the transcription factor binding sites (TFBS) influenced by these CpGs, we observed that most of the CpGs under-methylated in SZ-Clz potentially altered the binding of numerous transcription factors (TFs). Many of these TF have been previously associated with SZ. For instance, CpG1.3 methylation variability may impact the binding of ELK1, differentially expressed in SZ lymphoblastoid cells (58), FEV, linked to affective disorders (59), and SPDEF, which methylation moderates stress and substance abuse and its expression differs in the brain of patients with autism spectrum disorders and CTL (60). Similarly, CpG2.5 and CpG2.6 could affect the binding of EGR1, a stress response gene genetically associated with SZ (61).

Four, considering that many SZ-associated genetic variants have regulatory functions and that methylation is under local genetic control (62), we integrated genotypic variability in our analysis In the PFC, two variants (rs2208870 and rs12333117) along with LC1 scores or hypomethylated CpG units correlated with *NRN1* expression. HaploReg v4.2 data (63) showed that both variants alter TF binding affinity, with rs12333117 also affecting enhancer and promoter function in multiple tissues, including the brain.

Despite the insights gained into the effects of genetic variability and methylation patterns on *NRN1* expression in SZ, several limitations must be mentioned. Our relatively small sample size, although comparable to other studies, may not capture the full variability of the methylation landscape in SZ, limiting the generalizability of our findings. Additionally, the expression and methylation relationship with antipsychotics is two-way: antipsychotics can modulate methylation and thereby gene expression, but the unique methylation profile of an individual prior to treatment can also affect drug efficacy. Thus, without a group of never-treated patients, we cannot truly separate the effects of the disorder from those of the medication, but finding such post-mortem brain samples is challenging. Moreover, post-mortem samples also introduce variability due to tissue heterogeneity and differences in the cause of death, though we tried to account for this by including this last as a covariate. Finally, our cross-sectional study design limits our ability to assess changes in methylation over time, which is crucial for understanding SZ progression and treatment response. Moreover, establishing causality in such studies remains difficult. Therefore, further research with larger samples, longitudinal design, and advanced multi-omics approaches are needed to fully understand the epigenetic mechanisms underlying SZ.

In conclusion, we found that *NRN1* is significantly under-expressed in the PFC of patients with SZ treated with clozapine, with methylation levels correlating with its expression. Our study underscores distinct *NRN1* epigenetic patterns in the PFC and HIPP, highlighting the brain region specificity of SZ-related alterations. Additionally, incorporating genetic variability enhanced these correlations, emphasizing the importance of considering both genetic and epigenetic factors to fully understand the molecular mechanisms underlying SZ.

## Data Availability

All data produced in the present study are available upon reasonable request to the authors

## 6. ACKNOWLEDGEMENTS

This work was supported by: i) Fundación Alicia Koplowitz, ii) Acadèmia de les Ciències Mèdiques i de la Salut de Catalunya i de Balears. This study also received funding provided by: i) the Instituto de Salud Carlos III through a Miguel Servet contract to MF–V (CP20/00072), co-funded by European Regional Development Fund (ERDF)/European Social Fund “Investing in your future”; (i) the Acadèmia de les Ciències Mèdiques i de la Salut de Catalunya i de Balears through a contract to CA-P; iii) the Comissionat per a Universitats i Recerca del DIUE of the Generalitat de Catalunya, Agència de Gestió d’Ajuts Universitaris i de Recerca (AGAUR: 2021SGR1475); iv) the Basque Government (IT1512/22).

## 7. ETHICAL STATEMENTS

The study adhered to legal and ethical standards, with all samples collected at the Basque Institute of Legal Medicine (Bilbao, Spain) in accordance with Spanish national research policies and ethical guidelines for post-mortem brain studies in effect at the time of collection (Law 14/2007). Additional approval was obtained from the Research Ethics Committees of the Universidad del País Vasco/Euskal Herriko Unibertsitatea (UPV/EHU) in País Vasco, Spain (RD 1716/2011), and Germanes Hospitalaries in Cataluña, Spain (PR-2016-07).

## 9. AUTHORSHIP CONTRIBUTION STATEMENT

**Carmen Almodóvar-Payá:** Conceptualization, Data curation, Methodology, Investigation, Formal Analysis, Visualization, Writing – original draft, Writing – review & editing. **Marcos Moreno:** Data Curation, Methodology, Investigation, Formal Analysis, Writing – review & editing. **Maria Guardiola-Ripoll:** Data Curation, Investigation, Writing – review & editing. **Mariona Latorre-Guardia:** Methodology, Formal Analysis, Writing – review & editing. **Benito Morentin:** Data curation, Resources, Writing – review & editing. **Beatriz Garcia-Ruíz:** Investigation, Writing – review & editing. **Edith Pomarol-Clotet:** Resources, Funding acquisition, Writing – review & editing. **Luis F Callado:** Data Curation, Resources, Funding acquisition, Writing – review & editing. **Carme Gallego:** Conceptualization, Investigation, Methodology, Resources, Funding acquisition, Supervision, Writing – review & editing. **Mar Fatjó-Vilas:** Conceptualization, Methodology, Resources, Funding acquisition, Supervision, Writing – original draft, Writing – review & editing.

