## Supplementary material for "CLOZAPINE-RELATED BRAIN *NRN1* EXPRESSION PATTERNS ARE ASSOCIATED WITH METHYLATION AND GENETIC VARIANTS IN SCHIZOPHRENIA": Supplemetary Figures

### SUPPLEMENTARY FIGURES

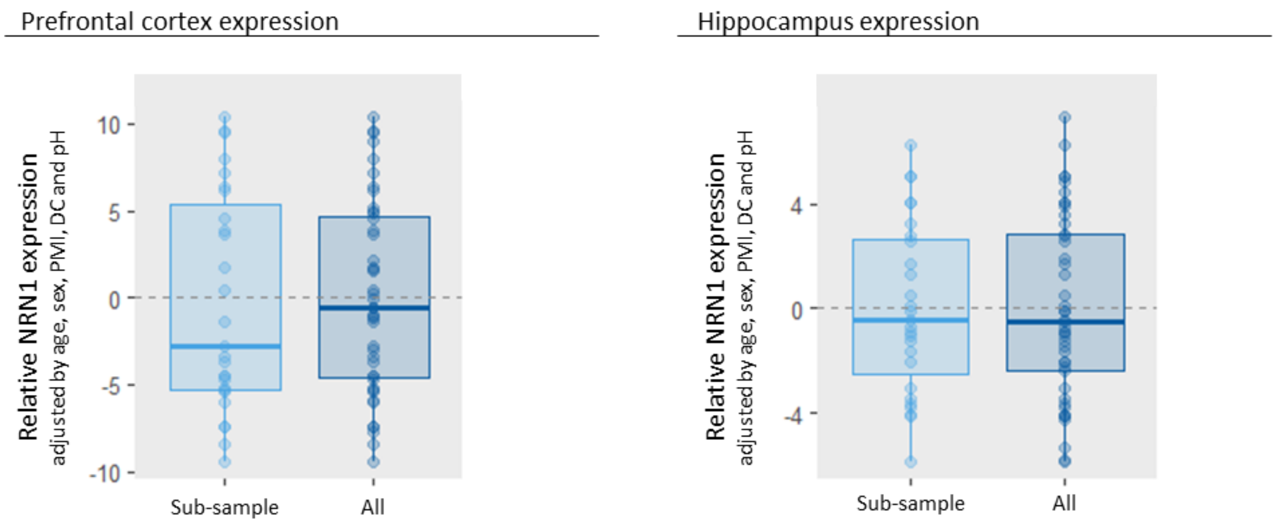


**Figure S1.** Box plots representing NRN1 gene expression levels ± 2 standard errors (se) in the general sample (All) and in the sub-sample use for methylation analyses for both brain regions. The expression of both genes was relativized to GAPDH and the values expressed are the residuals (adjusted by sex, age, post-mortem interval (PMI), death cause (DC) and pH).

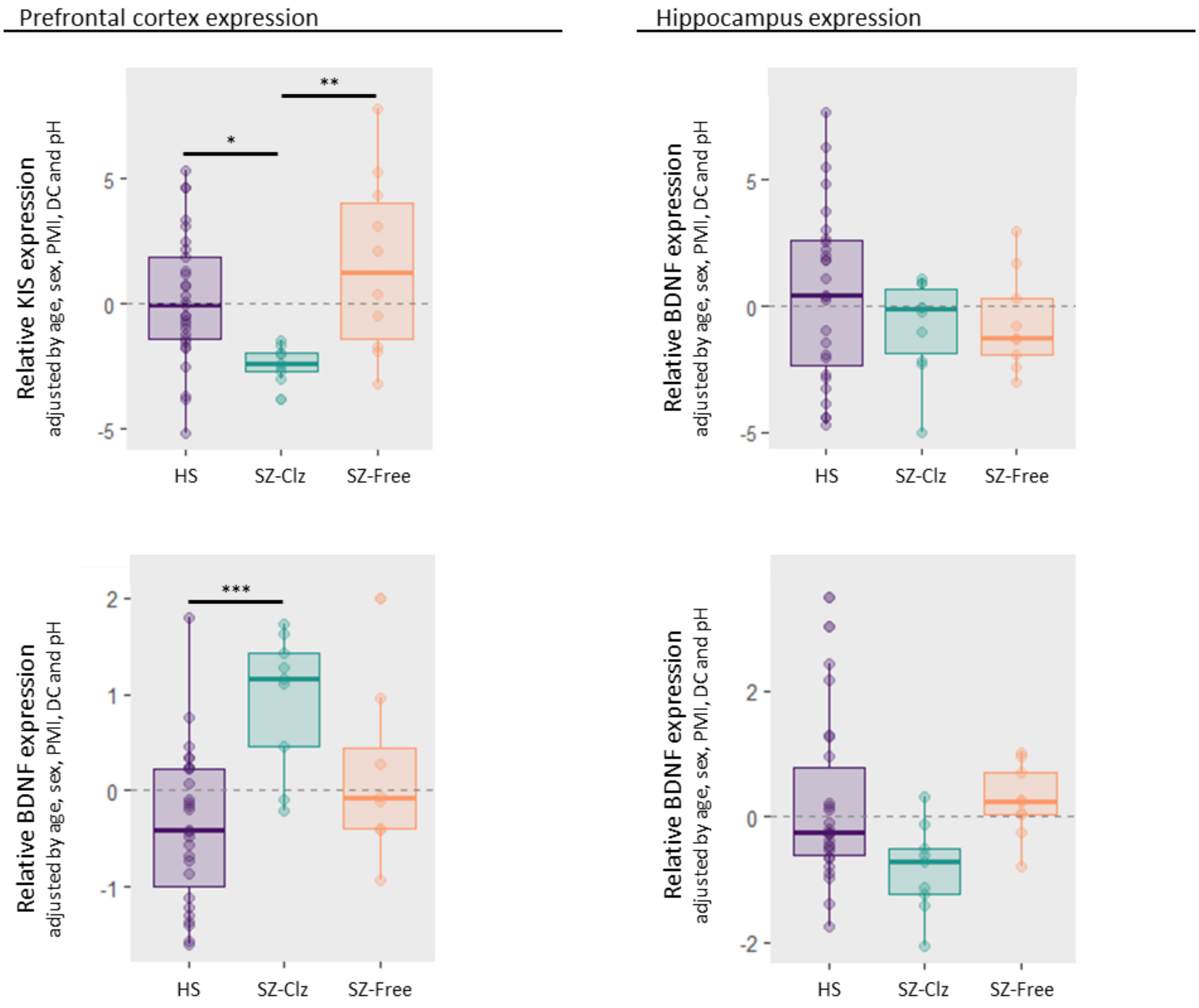


**Figure S2.** Box plots representing KIS and BDNF gene expression levels ± 2 standard errors (se) depicted according to the group, control subjects (CTL), schizophrenia patients with clozapine (SZ-Clz) or absence of antipsychotic (SZ-Free) in the blood at the time of death, for both brain regions: Prefrontal cortex (n=27, 9, and 9 respectively), and hippocampus (n=26, 9, and 10 respectively). The expression of both genes was relativized to GAPDH and the values expressed are the residuals (adjusted by sex, age, post-mortem interval (PMI), death cause (DC) and pH). Adjusted p-values from Tukey's test are represented as * $p<0.05$, ** $p<0.01$, *** $p<0.001$.

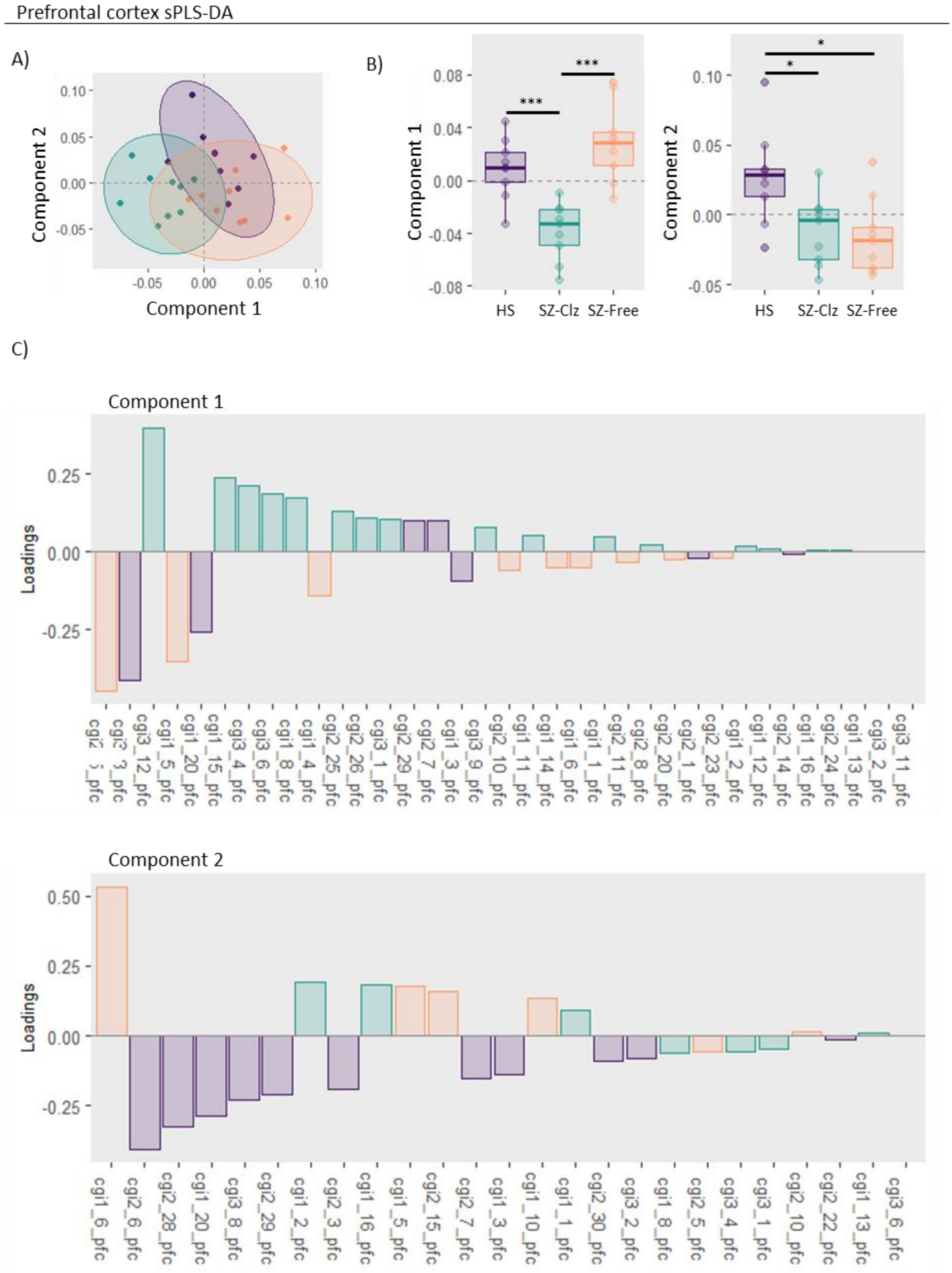


**Figure S3. A)** Scatter plot illustrating the distribution of samples projected into the space defined by the first two components derived from sparse Partial Least Square Discriminant Analysis (sPLS-DA) using *NRN1* methylation values from the prefrontal cortex (PFC). Each sample class is enclosed within 95% confidence ellipses. **B)** Boxplots representing methylation scores for the first and second latent components (LC) ± 2 standard errors (se) depicted according to the group. **C)** Bar plot illustrating the loading weights for each CpG unit included in the LC1 and LC2 with bars colored to indicate the group with higher mean methylation for each CpG unit. Control subjects (CTL) are depicted in purple, schizophrenia patients treated with clozapine (SZ-Clz) are shown in blue, and schizophrenia patients without antipsychotic treatment at the time of death (SZ-Free) are shown in orange. * p<0.05, ** p<0.01, *** p<0.001.

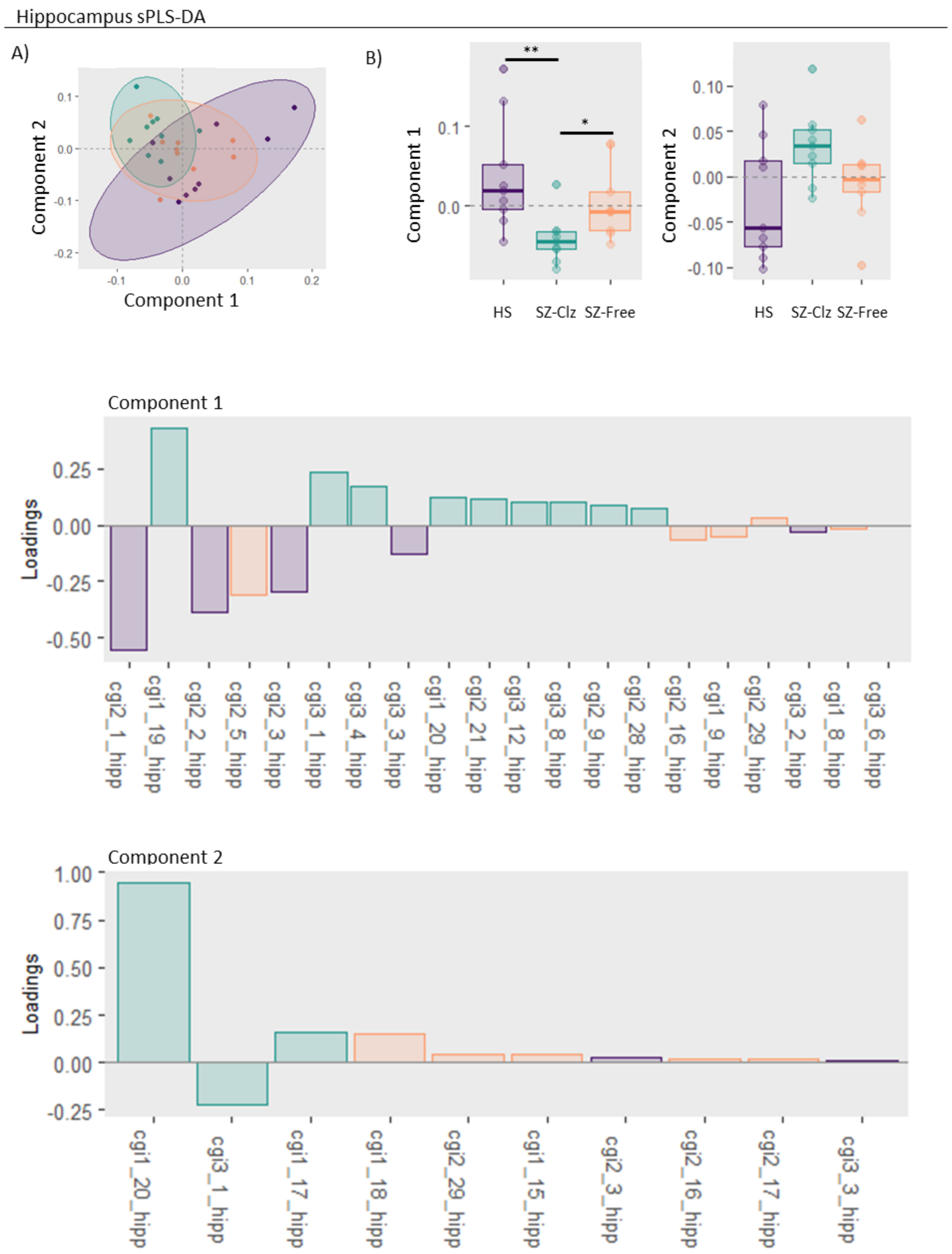


**Figure S4. A)** Scatter plot illustrating the distribution of samples projected into the space defined by the first two components derived from sparse Partial Least Square Discriminant Analysis (sPLS-DA) using *NRN1* methylation values from the hippocampus (HIPP). Each sample class is enclosed within 95% confidence ellipses. **B)** Boxplots representing methylation scores for the first and second latent components (LC) ± 2 standard errors (se) depicted according to the group. **C)** Bar plot illustrating the loading weights for each CpG unit included in the LC1 and LC2 with bars colored to indicate the group with higher mean methylation for each CpG unit. Control subjects (CTL) are depicted in purple, schizophrenia patients treated with clozapine (SZ-Clz) are shown in blue, and schizophrenia patients without antipsychotic treatment at the time of death (SZ-Free) are shown in orange. * p<0.05, ** p<0.01, *** p<0.001.

Prefrontal cortex

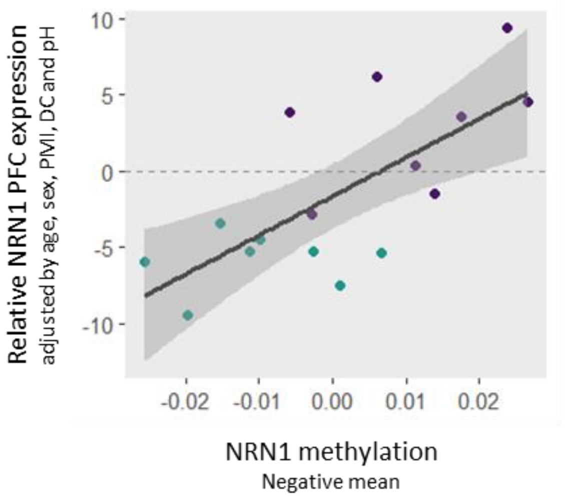


**Figure S5.** Scatter plots illustrating the correlation between the "negative mean" values, calculated using all CpG units included in latent component 1 derived from the sparse Partial Least Square Discriminant Analysis (sPLS-DA) of the prefrontal cortex (PFC), and NRN1 gene expression levels in the same region. These CpG units showed reduced methylation in schizophrenia patients treated with clozapine (SZ-Clz) in the blood at the time of death. NRN1 gene expression levels are shown with ±2 standard errors (SE). The expression of NRN1 was normalized to GAPDH, and the values are presented as residuals, adjusted for sex, age, post-mortem interval (PMI), cause of death (DC), and pH.
